# Environmental monitoring of Legionella in constructed water systems

**DOI:** 10.1101/2024.11.12.24317151

**Authors:** Samara Bin Salem, Abdullah Siddiqui, Premanandh Jagadeesan

## Abstract

Legionella is an opportunistic pathogen found in aquatic environments having profound health significance. Policy regulation mandates environmental monitoring for prevention and control of *Legionella* spp. in different types of water systems. The current study reports the trends of environmental monitoring of *Legionella* in constructed water system from the Emirate of Abu Dhabi. Sample collection and processing were performed as per standard procedures. A total of 8849 samples have been utilized during the 5 years study period of which 653 (7.4%) samples were positive for *Legionella* spp. The presence and frequency of different serogroups are presented. The relationship between residual chlorine levels and the presence of Legionella are discussed. The data indicates that most positive Legionella cases occur with residual chlorine levels of less than 0.2 mg/L while higher chlorine concentrations are associated with fewer Legionella-positive cases. In conclusion, environmental monitoring is very important to assess the trends of water quality for any remedial measures.

## Introduction

Legionella is an opportunistic pathogen found naturally in the environments. Presence of *Legionella* in aquatic environments is of profound health significance as the exposure may lead to Legionnaires’ disease, a severe form of pneumonia, and Pontiac fever, a milder illness resembling the flu. The disease can be fatal at times, particularly for immunocompromised individuals and people with chronic illnesses [1].

As a result, regulators across the globe have established management programs on an ad hoc basis or mandated routine monitoring by law. Guidelines have been developed to provide frameworks for risk assessment, management, and prevention strategies tailored to different types of water systems [2]. For instance, the World Health Organization (WHO), recommended water safety plan (WSP) which includes specifications for construction and commissioning of building water systems [3]. However, the guideline lacks threshold limits for Legionella unlike other microbial pathogens in water [4]. While guidelines are usually advisory and not necessarily prompt enforcement, the European Union Drinking Water Directive mandates monitoring of *Legionella* bacteria in all drinking water distribution systems, including large domestic water and industrial distribution systems. The parametric value of Legionella was <1000 CFU/L above which resampling should be performed and screened for *L. pneumophila* [5]. In the USA, there are no nationwide regulations for *Legionella* management [6].

Nevertheless, Center for disease control (CDC) has recommended a multifactorial approach as performance indicator interpretation. Accordingly, detection of up to 0.9 CFU/mL in a few of many tested locations within a water system is considered well controlled. The facility is considered as poorly controlled when more than 1CFU – 9.9 CFU/ml is found and above 10 CFU/ml is uncontrolled in potable water [7].

In Abu Dhabi, United Arab Emirates (UAE), the department of energy is the competing authority responsible for setting policy, standards and regulation for the energy sector which includes water. The regulation states a comprehensive risk assessment and periodic monitoring as effective means to ensure water safety which specifies a zero tolerant policy for wholesome water [8]. In the case of health care sector, all healthcare facilities are mandated to comply with water quality testing twice a year and notify Abu Dhabi Public Health Center (ADPHC) of any positive results. The current study aims to report the status of environmental monitoring of Legionella in constructed water system from the Emirate of Abu Dhabi.

## Materials and Methods

### Sample collection

Samples were collected in accordance with BS 7592:2022. Briefly, one liter of pre-flush samples was collected immediately after a tap or fitting is opened while dip samples were collected by immersing 1-L sterile container into a body of water depending on the sample source. All presterilized sample bottles contained 0.01% sodium thiosulfate to neutralize residual chlorine. The samples were labelled and transported in insulated containers to the laboratory for culture analysis as per ISO 11731-2: 2017.

### Sample Processing

Water samples were concentrated by membrane filtration using polycarbonate filters with a porosity of 0.2 µm. The filters were then transferred onto plates of the chosen culture medium selective for Legionella (BCYE Agar) and incubated at 37 ±1°C for 7-10 days. After the incubation period, morphologically characteristic colonies on the selective culture media were regarded as presumptive *Legionella* and were subjected to subculture to demonstrate their growth requirement for L-cysteine and iron (III) and species identification by LATEX agglutination. The confirmed number of *Legionella* were reported as colony forming units (CFU) of *Legionella* sp. or serogroups.

### Residual Chlorine

For the detection of residual chlorine, dpd (diethyl paraphenylene diamine) indicator test is applied. The colorimetric method measures the intensity of color produced when dpd reacts with residual chlorine. The intensity of the color produced is proportional to the concentration of the chlorine in the sample. The results are expressed as mg/l with two decimal points.

### Statistical Analysis

A descriptive statistical analysis was performed using Microsoft Excel. Non-parametric Mann–Whitney U test was conducted to establish the correlation between Legionella and residual chlorine (expressed in mg/L). The statistical results were interpreted at the level of significance p < 0.05.

## Results and discussion

Sampling and testing are the crucial step through which the effectiveness of water quality maintenance procedure is verified. Therefore, regulatory authorities enforce periodic monitoring of Legionella to assess the status in the Emirate of Abu Dhabi. While new rapid method such as real-time PCR analysis are best suited for well managed containment zones, culture is still regarded as the “gold standard” despite long incubation period as it provides concentration of cultivable *Legionella* which aids in risk management [9].

A total of 8849 samples have been utilized during the 5 years study period. The samples were taken from either taps or cistern of the respective collection location. Out of the 8849 samples collected, 653 (7.4%) samples were positive for *Legionella* spp. Further analysis revealed *L. pneumophila* serogroup 2-14 with a frequency of 19.6% while *L. pneumophila* serogroup 1 was found only in 2.8% of all positive samples. The number of samples analyzed during the study period and percentage of positive samples are presented in fig.1. It is evident from the results that *Legionella* persists in some sampling locations. Nevertheless, only 33 samples (0.4%) were in the range of 100-800 cfu/L while the remaining samples were between of 1-99 cfu/L (Table. 1). The actionable threshold of Legionella varies ranging from 10^2^ to 10^5^ cfu/L in potable water across Europe, UK and USA [10]. Since Abu Dhabi has adapted a zero tolerant policy, any presence of Legionella requires disinfection-based control measures to curb colonization. The infective dose of *Legionella* is a subject of debate [11] and hence such stringent measures are deemed necessary especially when there is no known safe level of *Legionella* in the water systems.

**Fig 1.**
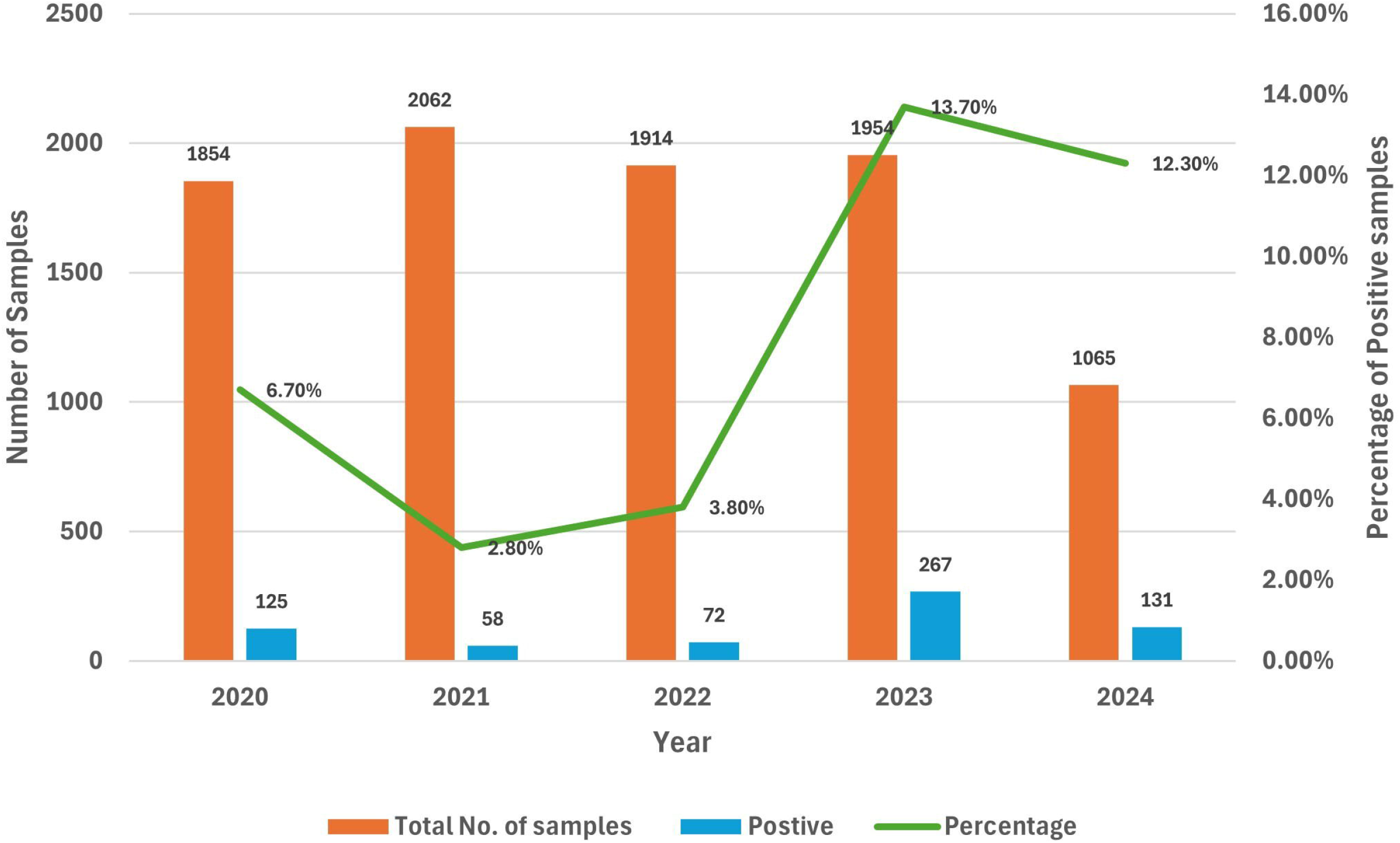

As far as the residual chlorine levels are concerned, there is a statistically significant difference between Legionella-positive and Legionella-negative samples (p-value < 0.05). The median chlorine level in Legionella-positive samples (0.1200 mg/L) is notably lower than in Legionella-negative samples (0.2260 mg/L). The analysis strongly suggests an inverse relationship between residual chlorine levels and the presence of Legionella in water samples. For instance, higher chlorine levels are associated with a lower likelihood of Legionella presence. The data indicates that most positive Legionella cases occur with residual chlorine levels less than 0.2 mg/L. As the chlorine concentration increases, the number of positive cases decreases, with the lowest number of cases in the category with chlorine levels of 0.5 mg/L (Fig. 2). This trend suggests that higher chlorine levels are associated with fewer Legionella-positive cases. The results signify the role of residual chlorine in the disinfection process and control of *Legionella* in constructed water bodies. The result from the study corroborates with previous studies on effective chlorine levels [12, 13] and reinstates the need of monitoring and maintaining the residual chlorine between 0.2-0.5 mg/L [8]. The stringent quality monitoring by both water and health care regulators mandating testing regime every 6 months has been a crucial tool to ensure the delivery of safe and good quality water to consumers.

**Fig 2.**
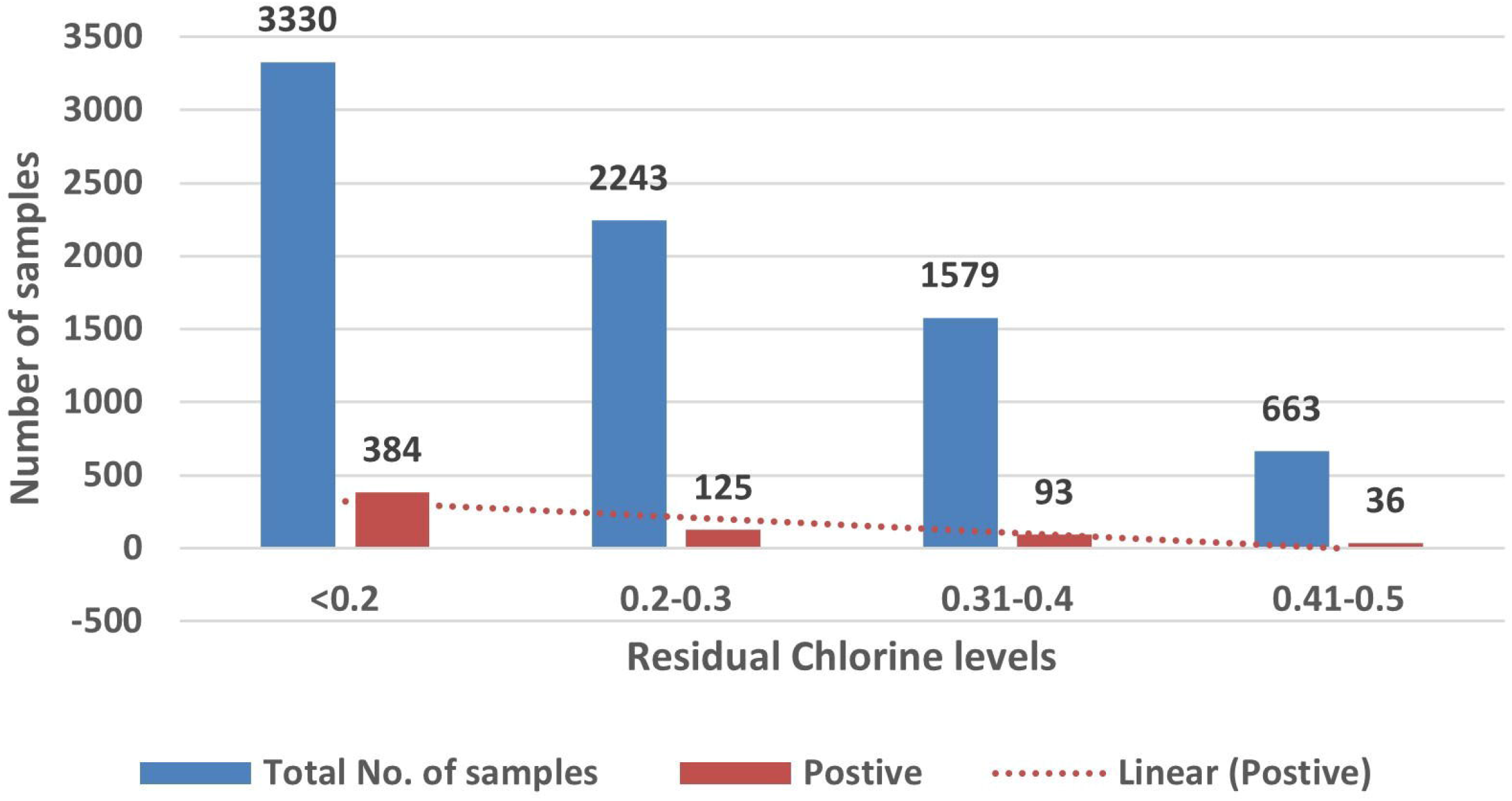

## Concluding remarks

A positivity rate of 7.4% has been observed over the study period with a surge in 2023 and 2024 emphasizing the impact of environmental monitoring of constructed water systems. This study further confirms the significance of maintaining residual chlorine between 0.2-0.5 mg/L as an effective means to control *Legionella* as recommended by the regulator. However, further investigation should reveal the resilience of these organism especially in areas where adequate concentration of chlorine is maintained.

## Data Availability

All data produced in the present work are contained in the manuscript

## Acknowledgments

The authors gratefully acknowledge the support and encouragement of the Abu Dhabi Quality and Conformity Council. Gratitude is also extended to the management of Central Testing Laboratories (CTL) for invaluable support.

