## Supplementary material for "Environmental monitoring of Legionella in constructed water systems": Table. 1

| **Legionella range (CFU/L)** | **Number of samples** | **Percentage** |
| --- | --- | --- |
| 1-10 | 241 | 2.7% |
| 11-99 | 379 | 4.3% |
| 100-800 | 33 | 0.4% |
| Negative | 8196 | 92.6% |

Table. 1*. Legionella* spp. range, and percentage of studied samples
